# SARS-CoV-2 RNA concentrations in primary municipal sewage sludge as a leading indicator of COVID-19 outbreak dynamics

**DOI:** 10.1101/2020.05.19.20105999

**Authors:** Jordan Peccia, Alessandro Zulli, Doug E. Brackney, Nathan D. Grubaugh, Edward H. Kaplan, Arnau Casanovas-Massana, Albert I. Ko, Amyn A. Malik, Dennis Wang, Mike Wang, Joshua L. Warren, Daniel M. Weinberger, Saad B. Omer

**Author notes:** Contributed equally to this work (Jordan Peccia, Alessandro Zulli, Doug E. Brackney). Corresponding authors: Jordan Peccia;: Saad Omer.

## Abstract

We report a time course of SARS-CoV-2 RNA concentrations in primary sewage sludge during the Spring COVID-19 outbreak in a northeastern U.S. metropolitan area. SARS-CoV-2 RNA was detected in all environmental samples, and when adjusted for the time lag, the virus RNA concentrations tracked the COVID-19 epidemiological curve. SARS-CoV-2 RNA concentrations were a leading indicator of community infection ahead of compiled COVID-19 testing data and local hospital admissions. Decisions to implement or relax public health measures and restrictions require timely information on outbreak dynamics in a community.

## Introduction

The most common metric followed to track the progression of the COVID-19 epidemic within communities is derived from testing symptomatic cases and evaluating the number of positive tests over time.^1^ However, tracking positive tests is a lagging indicator for an epidemic’s progression.^2, 3^ Testing is largely prompted by symptoms, which may take up to five days to present^4^, and individuals can shed virus prior to exhibiting symptoms. There is a pressing need for additional methods for early sentinel surveillance and near real-time estimations of community disease burden so that public health authorities may modulate and plan epidemic responses accordingly.

SARS-CoV-2 RNA is present in the stool of COVID-19 patients^5-7^ and has recently been documented in raw wastewater.^8-10^ Thus, monitoring raw wastewater (sewage) within a community’s collection system can potentially provide information on the prevalence and dynamics of infection for entire populations.^11^ When municipal raw wastewater discharges into treatment facilities, solids are settled and collected into a matrix called (primary) sewage sludge, which has been shown to contain a broad diversity of human viruses including commonly circulating coronavirus strains.^12^ Primary sludge provides a well-mixed and concentrated sample that may be advantageous for monitoring community shed SARS-CoV-2. As viral shedding can occur before cases are detected, we hypothesize that the time course of SARS-CoV-2 RNA concentrations in primary sewage sludge is a leading indicator of outbreak dynamics within a community served by the treatment plant.

## Results

During the COVID-19 outbreak from March 19, 2020 to May 1, 2020 in the New Haven, Connecticut (CT), USA metropolitan area, daily primary sludge samples were collected from the wastewater treatment facility which serves approximately 200,000 residents. SARS-CoV-2 viral RNA concentrations were quantitatively compared with local hospital admission data and community COVID-19 compiled testing data. SARS-CoV-2 viral RNA was detectable in all samples tested and ranged from 1.7 × 10^3^ virus RNA copies mL^-1^ to 4.6 × 10^5^ virus RNA copies mL^-1^. The lower concentration in this range corresponds to a qRT-PCR cycle threshold (CT) value of 38.75 and was used as a detection threshold for this method and sludge matrix. Overall, 96.5% of all CT values were less than 38 and values between 38 and 40 were reported as positive if detection occurred with virus nucleocapsid N1 and N2 primer sets and both replicates. Analysis of the human Ribonuclease P (RP) gene was utilized as a control, under the assumption that the RP gene content in primary sludge is constant over time and independent of SARS-CoV-2 RNA dynamics. Replicated samples demonstrated similar SARS-CoV-2 RNA concentration values, while comparisons between SARS-CoV-2 primer values with the human RP gene primer values indicate that dynamic temporal concentration changes in SARS-CoV-2 RNA were from actual change in virus concentration (**Figure 1A,B**). Concentration comparisons between replicates produced slopes of 0.97 (p<0.001) and 0.94 (p<0.001) for N1 and N2 primers, respectively, while comparing N and RP primer values resulted in a flat line, with slopes of −0.01(p=0.84) and −0.06 (p=0.25) for N1-RP and N2-RP, respectively (**Figure 1A,B**).

**Figure 1.**
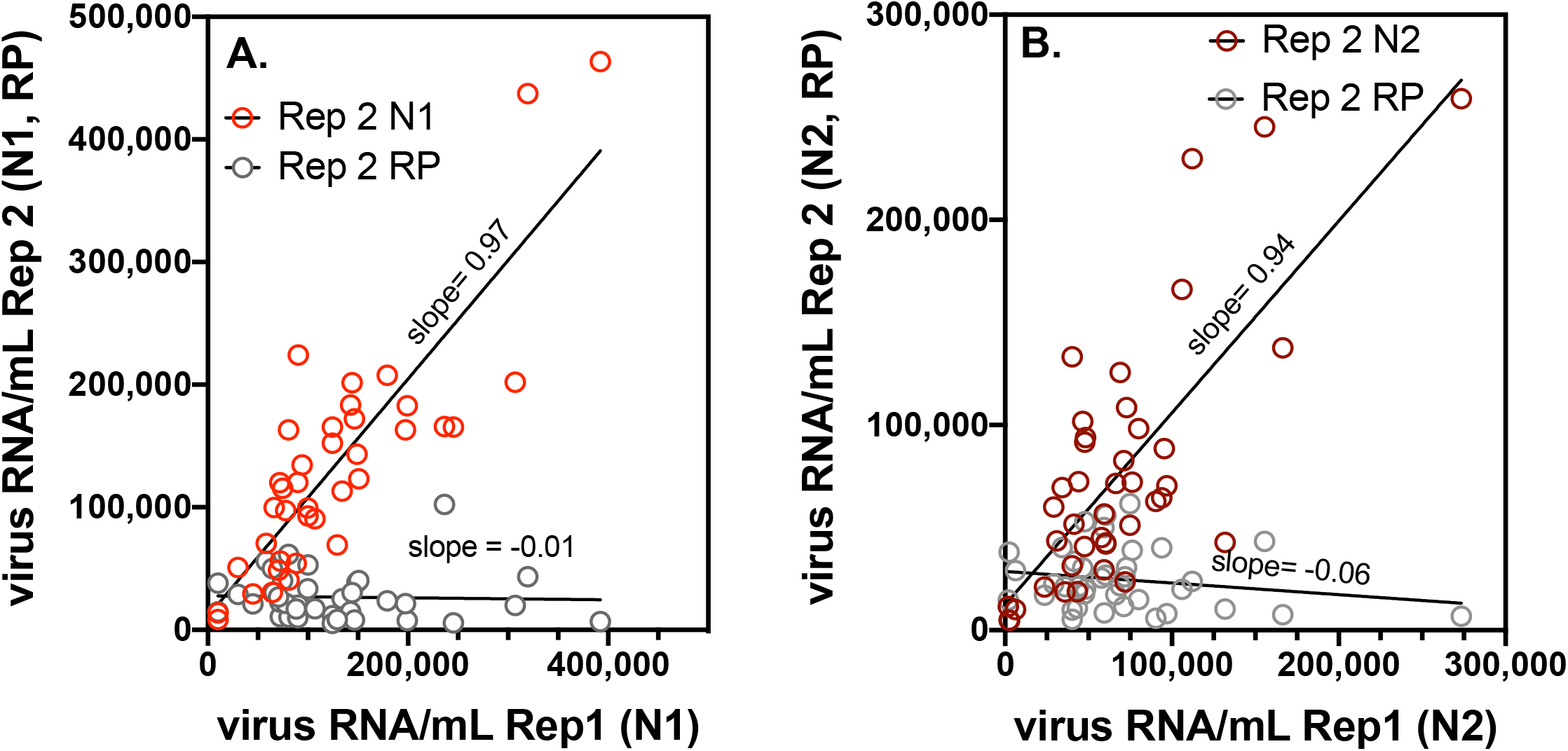
For the East Shore Water Pollution Abatement Facility, primary sludge SARS-CoV-2 concentration changes were observed during the COVID-19 outbreak, but changes in the human Ribonuclease P (RP) control gene were not observed**. (A)** Comparison of SARS-CoV-2 RNA concentrations between two replicate samples using the N1 primer set (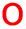), and comparison between N1 primer SARS-CoV-2 concentrations and human RP gene control concentrations (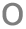); **(B)** Comparison of SARS-CoV-2 RNA concentration between two replicate samples using the N2 primer set (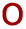), and comparison between N2 primer SARS-CoV-2 concentrations and human RP gene control concentrations (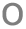);

Sampling during a New Haven COVID-19 outbreak suggests that these data may provide useful epidemiological insights. SARS-CoV-2 RNA sludge concentrations were quantitatively compared with data that are more commonly used to track the community progression of COVID-19 including hospital admissions (**Figure 2A,B**), the positive proportion of COVID-19 tests (by test date) for the four municipalities (New Haven, East Haven, Hamden, and Woodbridge, CT) served by the East Shore Water Pollution Abatement Facility (ESWPAF) (**Figure 2C,D**), and the publicly reported COVID-19 testing data based on reporting date for the four cities (**Figure 2E,F**). See **Figure 2G,H** for SARS-CoV-2 data. All four measures traced the initial wave of the SARS-CoV-2 outbreak in the New Haven metropolitan area; however, the sewage sludge SARS-CoV-2 RNA results showed a large increase in COVID-19 infections during the first week (March 19 to March 25) not observed in the hospitalization or testing data (**Figure 2**). Applying a distributed lag measurement error time series model allowed for estimating relationships between viral time series results and hospital admissions and testing data. By modeling the epidemiological time series as a function of the primary sludge SARS-CoV-2 RNA data across multiple daily lags (posterior means ± 90% credible intervals), viral RNA led hospital admissions by 1 to 4 days, proportion of positive tests by 0 to 6 days, and publicly reported testing data (reported date) by greater than 6 days with uncertainties in the reported testing estimates.

**Figure 2.**
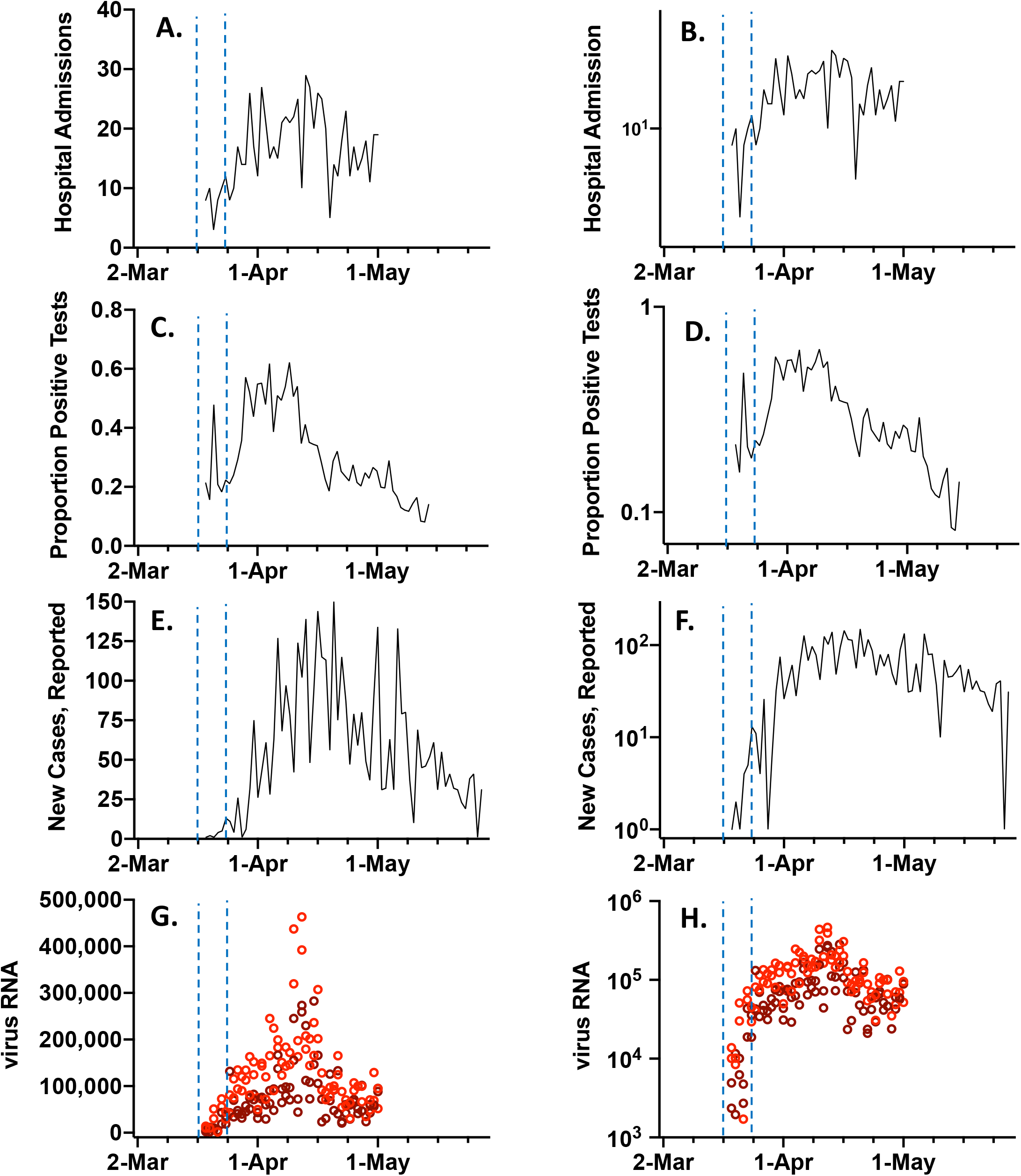
SARS-CoV-2 RNA concentration time course and other COVID-19 outbreak indicators on linear and log scales. All data represents the cities of New Haven, Hamden, East Haven, and Woodbridge, CT, USA, which are served by the East Shore Water Pollution Abatement Facility. The blue vertical dashed lines indicate the first week of analysis, March 19 to 25, 2020. **(A,B)** COVID-19 admissions to Yale New Haven Hospital for residents of the four cities (persons); **(C,D)** State of CT results for proportion of COVID-19 tests that were positive, reported by date tested; **(E,F)** Publicly available new COVID-19 cases by reporting date (persons); **(G,H)** Primary sludge SARS-CoV-2 RNA concentration (virus RNA gene copies mL^-1^ sludge) (N1 primers 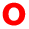; N2 primers 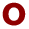).

## Discussion

We produced a SARS-CoV-2 RNA concentration time course in primary sewage sludge during a COVID-19 outbreak in the New Haven, CT metropolitan area. Approximately 200,000 people are served by the treatment facility and COVID-19 total documented cases by testing rose from less than 29 to 2,711 during the March 19 to May 1, 2020 surveillance period. Our results demonstrate: (1) the utility of SARS-CoV-2 primary sludge monitoring to accurately track outbreaks in a community and (2) primary sludge SARS-CoV-2 RNA concentrations can be a leading indicator over other commonly used epidemiology approaches including summarized COVID-19 test results and hospital admissions.

This study uniquely utilized primary sewage sludge instead of raw wastewater for virus RNA measurements. Sewage sludge is comprised of solids that are removed during primary sedimentation steps and typically gravity thickened. As a result, primary sludge has a greater solids content (2 to 5%) than raw wastewater (0.01 to 0.05%). Due to the elevated solids content and the high case load observed during the outbreak (~1,200 per 100,000 population), the concentrations of SARS-CoV-2 RNA reported here ranged from two to three orders of magnitude greater than raw wastewater SARS-CoV-2 values previously reported^8, 10, 13^. Monitoring primary sludge is broadly applicable. Wastewater treatment plants with primary and secondary treatment are standard in many regions of the world, and treatment facilities are expanding in urban areas of lower and middle-income countries.^14^ Within the US, approximately 16,000 treatment plants serve more than 250,000,000 people.^15^ The virus RNA time course tracked the COVID-19 case data, thus providing an approach to estimate the number of new cases from the relationship between new cases and SARS-CoV-2 RNA concentrations. However, these values are not presented here as we note that compiled COVID-19 testing is prompted by symptoms, thus typically excluding asymptomatic or mild infections, and underestimating the true number of cases.^16^

SARS-CoV-2 RNA concentrations in sewage sludge were a leading indicator of community outbreak dynamics over hospitalization and compiled COVID-19 testing data. The time from patients shedding to detection in primary sewage is typically on the order of hours, including a 2 to 4 hour conveyance to a wastewater facility, and less than 5 hour residence time in primary sedimentation equipment. By comparison, delays associated with the onset of COVID-19 symptoms (which prompt testing) and estimated times to hospitalization can be greater than 5 days. In addition, the COVID-19 epidemic arrived in the New Haven metropolitan area in early March, 2020 when testing capacity, access, and time to results were all under development during the beginning of the outbreak.

In conclusion, the SARS-CoV-2 RNA concentrations in primary sewage sludge were tracked throughout a COVID-19 epidemic and compared with traditional outbreak epidemiological indicators. SARS-CoV-2 RNA concentration in primary sludge followed the epidemiology curves established by compiled COVID-19 testing data and hospital admissions, but was a leading indicator. Our study could have substantial policy implications. Jurisdictions can use primary sludge SARS-CoV-2 concentrations to preempt community outbreak dynamics or provide an additional basis for easing restrictions, especially when there are limitations in clinical testing. Raw wastewater and sludge-based surveillance is particularly useful for low and middle-income countries where clinical testing capacity is limited.

## Methods

### Sample collection

Primary sewage sludge was collected from the East Shore Water Pollution Abatement Facility (ESWPAF) in New Haven, CT, USA. Samples were taken daily from March 19 to May 1, 2020 between 8 and 10 am EDT and stored at −80°C prior to analysis. The initial sampling dates were prior to widespread individual testing in the region, and prior to the start of stay at home restrictions implemented throughout the State of Connecticut, USA (**Figure 1A**). From the sampling start and end dates, cities served by the ESWPAF experienced greater than 85 times increase in confirmed COVID-19 cases (by reported testing) from less than 29 cases to 2,711.^17^ The plant serves an estimated population of 200,000 people with average treated flows of 1.75 m^3^/s. Sludge collected from ESWPAF is primary sludge, sampled at the outlet of a gravity thickener, ranging in solids content from 2.6% to 5%.

### Viral RNA quantitative testing

To quantify SARS-CoV-2 virus RNA concentrations in primary sludge, 2.5 mL of well mixed sludge were added directly to a commercial kit optimized for isolation of total RNA from soil (RNeasey PowerSoil Total RNA kit, Qiagen). Isolated RNA pellets were dissolved in 50 μL of ribonuclease free water and total RNA measured by spectrophotometry (NanoDrop, ThermoFisher Scientific). SARS-CoV-2 RNA was quantified by one-step quantitative reverse transcriptase-polymerase chain reaction (qRT-PCR) using the U.S. Center for Disease Control N1 and N2 primers sets.^18^ For control, analysis was also conducted for the human Ribonuclease P (RP) gene. Samples were analyzed using the Bio-Rad iTaq Universal Probes One-Step kit in 20 μL reactions run at 50°C for 10 min, 95°C for 1 min followed by 40 cycles of 95°C for 10 s and 60°C for 30 s per the manufacturer’s recommendations. SARS-CoV-2 RNA concentrations were determined using a standard curve as previously described^18^ and presented as virus RNA copies. The SARS-CoV-2 concentration results were normalized to the total RNA extracted to reduce day to day variations in treatment plant flow, sludge solids content, and RNA extraction efficiency. All samples were diluted 5x for use as template to ensure the removal of inhibition. Performing qRT-PCR on undiluted and 5x diluted samples typically resulted in the expected 5x decrease in concentration. Sewage sludge from 2018 was used as a control and no SARS-CoV-2 detection was observed from either N1 or N2 primers. These control sludges were consistently positive for the human RP gene.

### Epidemiological Analysis

Daily COVID-19 admissions to the Yale New Haven Hospital were compiled from hospital records upon request from the hospital’s Joint Data Analytics Team for the New Haven Health System and confirmed by laboratory testing. Hospital admissions only include residents from the towns served by the ESWPAF (New Haven, East Haven, Hamden, and Woodbridge CT). The proportion of tests of residents from the four cities that were positive for COVID-19 and reported by test date were supplied through a data request to the CT Department of Public Health. Publicly available numbers of laboratory-confirmed COVID-19 cases by reporting date in the towns served by the ESWPAF (New Haven, East Haven, Hamden, and Woodbridge CT) were compiled from daily reports published by the CT Department of Public Health.^17^

Estimation of primary sludge as a leading indicator was performed using a distributed lag measurement error time series model. This analysis was carried out in the Bayesian framework, allowing us to correctly characterize multiple sources of uncertainty when estimating the lagged associations of interest. See citation^19^ for a more in-depth description of this analysis.

## Data Availability

Once published, data will be available on the COVID tracker website.

https://covidtrackerct.com/

## Acknowledgements

We wish to thank the East Shore Water Pollution Abatement Facility, New Haven, CT, USA for primary sludge sampling assistance. AZ is supported by the Yale Institute of Global Health and NDG is supported by a gift from the Huffman Family Donor Advised Fund. The authors thank Kimberly Yousey-Hindes and Paula Clogher for providing the data on proportion positive tests.

